# Deep Learning Phenotyping of Tricuspid Regurgitation for Automated High Throughput Assessment of Transthoracic Echocardiography

**DOI:** 10.1101/2024.06.22.24309332

**Authors:** Amey Vrudhula, Milos Vukadinovic, Christiane Haeffle, Alan C. Kwan, Daniel Berman, David Liang, Robert Siegel, Susan Cheng, David Ouyang

## Abstract

**Background and Aims:** Diagnosis of tricuspid regurgitation (TR) requires careful expert evaluation. This study developed an automated deep learning pipeline for assessing TR from transthoracic echocardiography.

**Methods:** An automated deep learning workflow was developed using 47,312 studies (2,079,898 videos) from Cedars-Sinai Medical Center (CSMC) between 2011 and 2021. The pipeline was tested on a temporally distinct test set of 2,462 studies (108,138 videos) obtained in 2022 at CSMC and a geographically distinct cohort of 5,549 studies (278,377 videos) from Stanford Healthcare (SHC).

**Results:** In the CSMC test dataset, the view classifier demonstrated an AUC of 1.000 (0.999 – 1.000) and identified at least one A4C video with colour Doppler across the tricuspid valve in 2,410 of 2,462 studies with a sensitivity of 0.975 (0.968-0.982) and a specificity of 1.000 (1.00-1.000). In the CSMC test cohort, moderate-or-severe TR was detected with an AUC of 0.928 (0.913 - 0.943) and severe TR was detected with an AUC of 0.956 (0.940 - 0.969). In the SHC cohort, the view classifier correctly identified at least one TR colour Doppler video in 5,268 of the 5,549 studies, resulting in an AUC of 0.999 (0.998 – 0.999), a sensitivity of 0.949 (0.944 - 0.955) and specificity of 0.999 (0.999 – 0.999). The AI model detected moderate-or-severe TR with an AUC of 0.951 (0.938 - 0.962) and severe TR with an AUC of 0.980 (0.966 - 0.988).

**Conclusions:** We developed an automated pipeline to identify clinically significant TR with excellent performance. This approach carries potential for automated TR detection and stratification for surveillance and screening.

**Structured Graphical Abstract:** Computer Vision Based Tricuspid Regurgitation (TR) Detection: An automated deep learning pipeline was trained to stratify tricuspid regurgitation severity using large-scale data in the form of A4C TTE videos with colour Doppler across the tricuspid valve. The pipeline generalized across two geographically distinct test sets from CSMC and SHC, demonstrating the pipeline’s ability to detect clinically significant TR using single-view TTE videos with Doppler information. These results open the door to potential TR point-of-care screening.

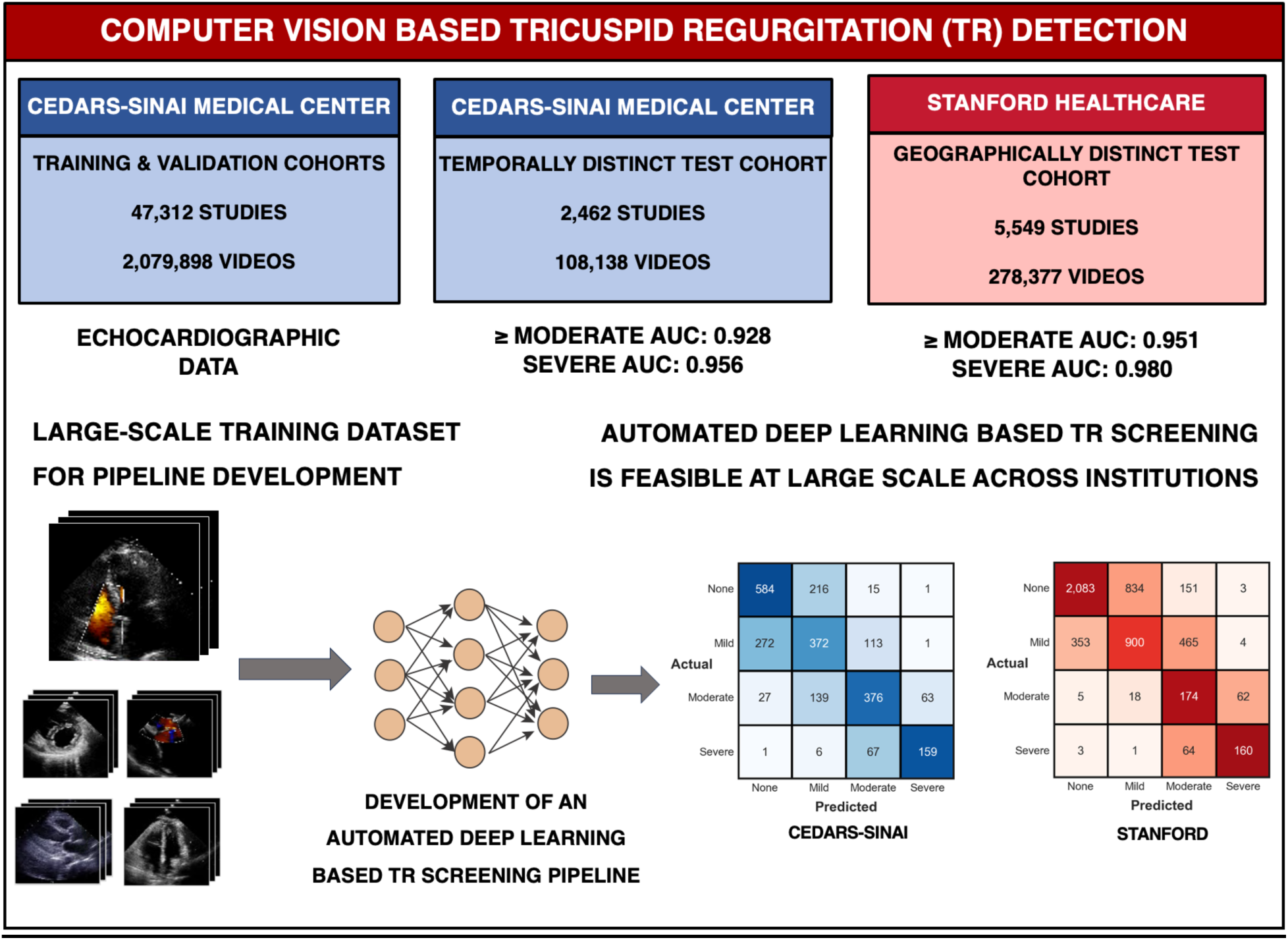

**Key Question:** Can an automated deep learning model assess tricuspid regurgitation severity from echocardiography?

**Key Finding:** We developed and validated an automated tricuspid regurgitation detection algorithm pipeline across two healthcare systems with high volume echocardiography labs. The algorithm correctly identifies apical-4-chamber view videos with colour Doppler across the tricuspid valve and grades clinically significant TR with strong agreement to expert clinical readers.

**Take Home message:** A deep learning pipeline could automate TR screening, facilitating reproducible accurate assessment of TR severity, allowing rapid triage or re-review and expand access in low-resource or primary care settings.

## Introduction

Accurate and reliable assessment of tricuspid regurgitation (TR) severity remains an ongoing challenge. Once considered a benign consequence of co-existing heart disease, tricuspid regurgitation has received more recent recognition as an independent risk factor for morbidity and mortality^1–3^. While recent findings highlight the need for earlier diagnosis and monitoring, early TR rarely presents with symptoms in the absence of co-existing cardiac disease. With high temporal resolution, transthoracic echocardiography is the most common test of choice for characterizing TR, however accurate diagnosis requires expert assessment as there is significant intra-observer variability^4,5^. As new therapeutic options for treating TR like percutaneous repair emerge, early and accurate diagnosis of tricuspid regurgitation continues to become more important.

Recent advances in computer vision and artificial intelligence (AI) have enabled precision phenotyping of structure and function in cardiac ultrasound^6^. AI applied to echocardiography can precisely estimate wall thickness^7^, assess mitral regurgitation severity^8^, and left ventricular ejection fraction (LVEF)^9,10^, as well as detect cardiac amyloidosis^7,11^, HCM^12^, and diastolic dysfunction^13^. Application of deep learning for tricuspid regurgitation has lagged behind, with most machine learning approaches using structured tabular data to characterize and prognosticate TR rather than evaluating the underlying images themselves^14,15,16,17^. AI guidance has been developed for both image acquisition and interpretation^9,18^, and given the increasing prevalence of TR in an aging population with co-morbid heart failure, AI could aid in TR screening and surveillance^19–23^.

In the present study, we developed and evaluated a deep learning pipeline to detect and assess TR severity from transthoracic echocardiogram studies. Automating the entire process of view selection, identification of tricuspid regurgitation by colour Doppler, and assessment of severity, we hypothesized that a deep learning approach could assess TR severity with high-throughput automation. This pipeline was evaluated with data from two geographically distinct sites, including temporally distinct test cohort distinct from the training and validation cohorts **(Figure 1)**. Combined with other echocardiography AI algorithms, like those enabling novices to obtain point of care echocardiographic images^18^, such an approach could be used for serial surveillance and screening of tricuspid regurgitation.

**Figure 1:**
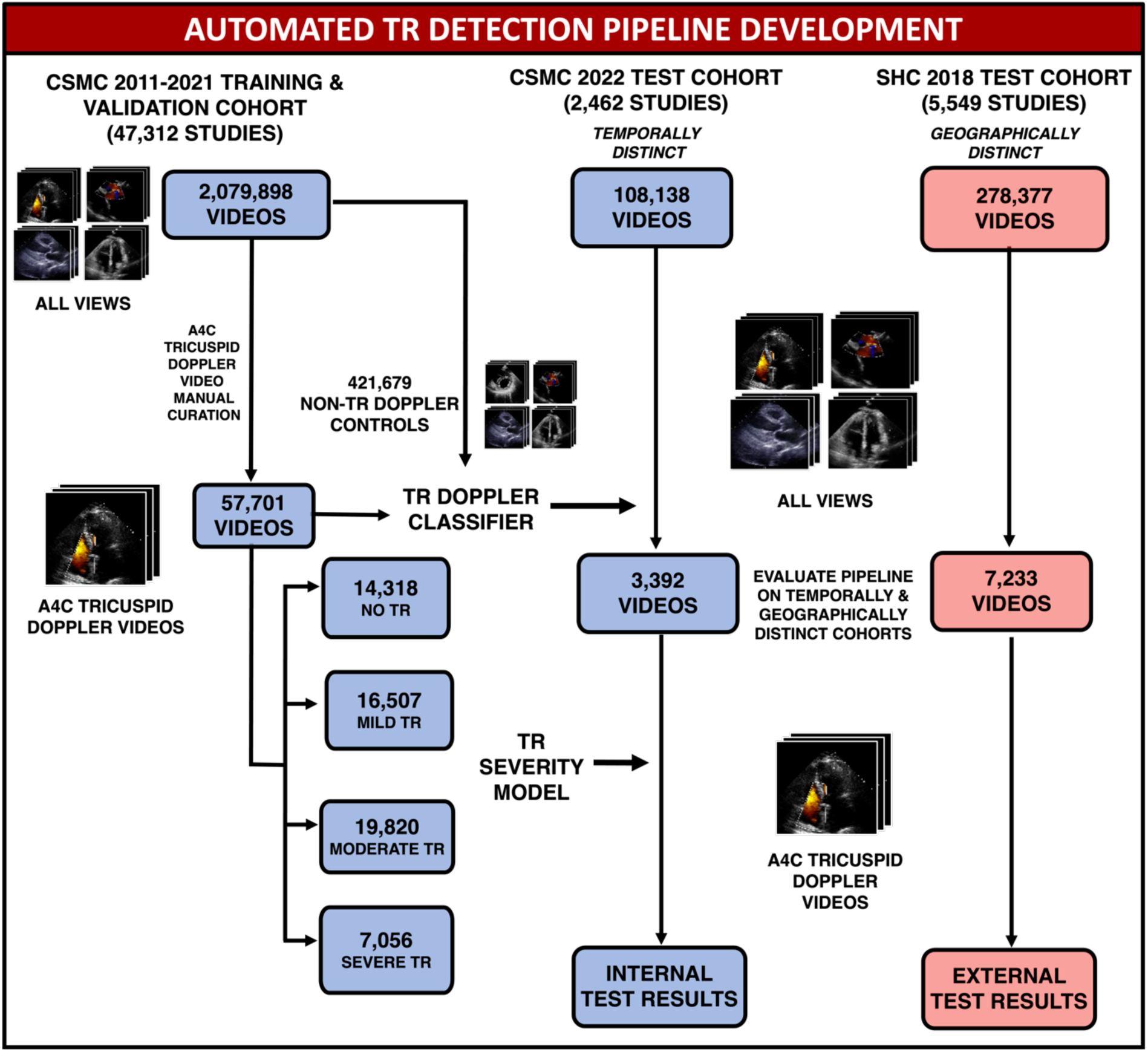
CSMC and Stanford Dataset Isolation. - 57,701 A4C videos were manually curated from 47,312 studies with varying TR severity and used to train deep learning models for view classification and TR severity stratification. A pipeline consisting of these models was then benchmarked on a temporally distinct cohort of 2,462 studies from CSMC and a geographically distinct cohort of 5,549 studies from SHC.

## Methods

### Study Population and Data Source

***Cedars-Sinai Medical Center (CSMC) Cohort:*** Transthoracic echocardiography (TTE) studies at Cedars-Sinai Medical Center (CSMC) between October 04, 2011 and December 31, 2021 were used to train our deep learning pipeline (‘EchoNet-TR’) for high-throughput TR identification and grading. Studies were initially sourced from Digital Imaging and Communications in Medicine (DICOM) files and underwent de-identification, view classification, and pre-processing into AVI files as previously described^8^, yielding 2,079,898 videos from 47,312 studies from 31,708 patients. From these videos, 57,701 A4C videos with colour Doppler across the tricuspid valve were manually curated and used to train a deep learning pipeline for TR phenotyping.

Studies were randomly split on a patient level into train (95%) and internal validation (5%) cohorts to train deep neural networks for TR phenotyping.^24^ Identical patient-level splits were maintained training both the TR severity and view classification models. The trained models were evaluated serially as a single pipeline on a held-out temporal test set of 2,462 TTE studies (101,455 videos) from 2,170 patients receiving care at CSMC between January 01, 2022 and June 04, 2022. Patients in the training and validation sets were excluded from the training set.

***Stanford Healthcare (SHC) Cohort:*** The pipeline was evaluated on 5,549 studies (containing a total of 278,377 videos) from SHC’s high-volume echocardiography lab. The automated view classification pipeline was compared with manual curation of videos within those studies to evaluate specificity. All videos identified by the view classifier were used for downstream TR severity model validation. Model output was compared with TR severity determined by expert cardiologists from the clinical reports. This study was approved by the Institutional Review Boards at Cedars-Sinai Medical Center and Stanford Healthcare. Informed consent was waived as the study involved secondary analysis of existing data without patient contact.

### AI Model Training

The model pipeline consisted of a view classifier capable of detecting A4C videos with colour Doppler across the tricuspid valve from full echocardiographic studies and a TR severity classification model. The PyTorch Lightning deep learning framework was used to train deep learning models. Video-based convolutional neural networks of the R2+1D architecture were used for view classification and TR severity assessment.^25^ The view classification model was initialized with random weights while the TR severity model was initialized with weights from Echo-Net Dynamic^9^. Both models were trained using a cross-entropy loss function for up to 100 epochs, an ADAM optimizer, an initial learning rate of 1e-2, and a batch size of 24 on an NVIDIA RTX 3090 GPU. Early stopping was performed based on the validation loss.

The view classifier was trained using the 57,701 manually curated A4C videos with colour Doppler across the tricuspid valve as cases and 421,679 controls from the same studies. Controls consisted of any videos that were not A4C videos with colour Doppler across the tricuspid valve and included videos of both A4C and other echocardiographic views (both with and without colour Doppler information). The TR severity model was trained using 57,701 manually curated videos, which consisted of 14,318 videos with no TR, 16,507 videos with mild TR, 19,820 videos with moderate TR, and 7,056 videos with severe TR. TR severity for each study was determined based on the clinical echocardiographic reports from CSMC’s high-volume echocardiography lab, where severity was assessed in accordance with ASE guidelines.^26^ When TR was characterized as an intermediate category (ie. “trace to mild” or “mild to moderate” or “moderate to severe”), videos were placed in the more severe category. Studies with concomitant tricuspid stenosis, prosthetic valves, and heart failure were also included in both training and validation datasets.

### Statistical Analysis

The pipeline was then evaluated on two test sets not seen during model training: 2,462 studies obtained at CSMC in 2022 (temporally distinct from the training and validation studies and with no patient overlap) and 5,549 studies obtained at SHC in 2018. This process is summarized in **Figure 1**. Confusion matrices and area under the receiver operating characteristic curve (AUC) were used to assess model performance, and statistics related to TR model performance were calculated on a study level. When a study had more than one A4C video with Doppler information across the tricuspid valve, the video that resulted in the greatest predicted TR severity was used for analysis. When multiple videos resulted in the same maximal predicted severity, the video with the highest prediction probability for the given level of TR severity was used. In both the internal and external test sets, AUC, F1-score, recall (sensitivity), positive predictive value (PPV), and negative predictive value (NPV) were calculated for clinically significant TR, which was defined as greater than moderate TR and severe TR. AUC was also calculated for relevant subsets in the CSMC test cohort. Statistical analysis was performed in Python (version 3.8.0). Confidence intervals were computed via bootstrapping with 10,000 samples. Reporting of study results is consistent with guidelines put forth by CONSORT-AI.^27,28^

Subgroup analysis was conducted to assess model performance in patients with different ranges of right and left ventricular ejection fraction, pulmonary artery pressure, associated co-morbidities (≥ mild right atrial dilation, ≥ mild mitral regurgitation, ≥ moderate aortic stenosis, ≥ moderate aortic regurgitation), study characteristics, associated co-morbidities and other clinical characteristics. Echocardiogram study quality was determined by clinicians and extracted from the clinical report. Studies where clinicians commented on technical difficulty, poor study quality, or where one or more major cardiac structures (left ventricle, right ventricle, pulmonary artery, etc.) were not well visualized were classified as technically difficult.

### Model Explainability

Features identified by the TR severity model were evaluated using saliency mapping, generated using the Integrated Gradients method.^29^ This method generated a heatmap for every frame of the video, summarized as a final 2-dimensional heatmap generated by using the maximum value along the temporal axis for each pixel location in the video. Pixels brighter in intensity and closer to yellow were more salient to model predictions, while those darker in colour were less important to the model’s final prediction. When assessing videos with no TR, heatmaps were obtained by taking the maximum of saliency maps for the moderate and severe class output neurons for each pixel location.

## Results

### Study Population

Patient characteristics are shown in **Tables 1 & 2**. In the CSMC training, validation, and test sets, patients had similar characteristics. Meanwhile, the CSMC and SHC test cohorts differed. The SHC test cohort contained lower numbers of studies from patients with moderate TR (5.0% vs. 25.0%) and severe TR (4.4% vs. 10.0%). The SHC cohort also had a lower proportion of videos from Black patients (4.5 % vs. 14.5%), and a higher proportion of videos from Asian patients (25.2% vs. 9.8%).

**Table 1.**

|  | Derivation Cohorts |  | Test Cohorts |  |
| --- | --- | --- | --- | --- |
|  | Train | Val | CSMC Test Cohort | SHC Test Cohort |
| **Patients** | 30,125 (88.92) | 1,583 (4.67) | 2,170 (6.41) | 5,014 |
| **Studies** | 44,908 | 2,404 | 2,462 | 5,549 |
| **Videos** | 453,787 | 24,484 | 101,415 | 278,377 |
| **Male** | 23,552 (53.0) | 1,282 (53.3) | 1,286 (52.2) | 1,057 (52.7) |
| **HTN** | 27,743 (61.8) | 1,477 (61.4) | 1,295 (52.6) | 953 (47.6) |
| **CAD** | 18,983 (42.3) | 999 (41.6) | 903 (36.7) | 721 (36.0) |
| **AF** | 14,612 (32.5) | 856 (35.6) | 515 (20.9) | 550 (27.4) |
| **EF** | 56.9 (15.4) | 57.0 (15.1) | 57.2 (15.1) | 57.4 (11.0)* |
| **LAVI** | 35.5 (15.7) | 36.1 (16.0) | 31.8 (15.5) | N/A |
| **TR Severity** |  |  |  |  |
| Control | 12297 (27.4) | 627 (26.1) | 829 (33.7) | 3,223 (58.1) |
| Mild | 13286 (29.6) | 710 (29.5) | 770 (31.3) | 1,808 (32.6) |
| Moderate | 14363 (32.0) | 785 (32.7) | 617 (25.1) | 275 (5.0) |
| Severe | 4962 (11.0) | 282 (11.7) | 246 (10.0) | 243 (4.4) |
| **Race** |  |  |  |  |
| White | 31,117 (69.3) | 1,698 (70.6) | 1,559 (63.3) | 1,097 (54.8) |
| Black | 6,004 (13.4) | 288 (12.0) | 358 (14.5) | 90 (4.5) |
| Asian | 3,459 (7.7) | 192 (8.0) | 241 (9.8) | 504 (25.2) |
| Other | 4,328 (9.6) | 226 (9.4) | 304 (12.3) | 311 (15.5) |

*2003 studies in the SHC test cohort had information on comorbidities, demographics, and EF.*

**Table 2.**

|  | Derivation Cohorts |  | Test Cohorts |  |
| --- | --- | --- | --- | --- |
|  | Train | Val | CSMC Test Cohort | SHC Test Cohort |
| **Patients** | 30125 (88.92) | 1583 (4.67) | 2,132 (6.41) | 4,798 |
| **Studies** | 44,908 | 2,404 | 2,412 | 5,280 |
| **Videos** | 453,787 | 24,484 | 3,392 | 7,233 |
| **Male** | 23,552 (53.0) | 1,282 () | 1263 () | 1,057 (52.7) |
| **HTN** | 27,743 (61.8) | 1,477 (61.4) | 1271 (52.7) | 898 (47.3) * |
| **CAD** | 18,983 (42.3) | 999 (41.6) | 884 (36.7) | 674 (35.5) * |
| **AF** | 14,612 (32.5) | 856 (35.6) | 499 (20.7) | 520 (27.4) * |
| **EF** | 56.9 (15.4) | 57.0 (15.1) | 57.2 (16.9) | 57.5 (10.8)* |
| EF ≥ 35 | 39,294 (89.5) | 2,111 (90.0) | 2105 (89.1) | 1,796 (93.7) * |
| EF < 35 | 4,616 (10.5) | 236 (10.0) | 255 (10.8) | 120 (6.3) * |
| EF ≥ 50 | 35,287 (80.4) | 1,889 (80.5) | 1924 (81.5) | 1,585 (82.7) * |
| EF < 50 | 8623 (19.6) | 458 (19.5) | 436 (18.5) | 331 (17.2) * |
| **LAVI** | 35.5 (15.7) | 36.1 (16.0) | 31.8 (15.3) | N/A |
| **TR Severity** |  |  |  |  |
| **Control** | 12297 (27.4) | 627 (26.1) | 829 (33.7) | 816 (33.8) |
| **Mild** | 13286 (29.6) | 710 (29.5) | 770 (31.3) | 758 (31.4) |
| **Moderate** | 14363 (32.0) | 785 (32.7) | 617 (25.1) | 605 (25.1) |
| **Severe** | 4962 (11.0) | 282 (11.7) | 246 (10.0) | 233 (9.7) |
| **Race** |  |  |  |  |
| **White** | 31,117 (69.3) | 1,698 (70.6) | 1,559 (63.3) | 1,097 (54.8) |
| **Black** | 6,004 (13.4) | 288 (12.0) | 358 (14.5) | 90 (4.5) |
| **Asian** | 3,459 (7.7) | 192 (8.0) | 241 (9.8) | 504 (25.2) |
| **Other** | 4,328 (9.6) | 226 (9.4) | 304 (12.3) | 311 (15.5) |
| **RV Dysfunction** |  |  |  |  |
| Mildly Depressed | 7,433 (13.6) | 374 (12.8) | 415 (12.3) | N/A |
| Moderately/Severely Depressed | 6,027 (11.0) | 301 (10.3) | 409 (12.1) | N/A |
| **Pulmonary Artery Pressure** |  |  |  |  |
| PA Pressure > 35 mm Hg | 17,646 (44.6) | 976 (46.1) | 822 (39.7) | N/A |
| PA Pressure ≤ 35 mm Hg | 21,866 (55.4) | 1,140 (53.9) | 1,245(60.2) | N/A |
| PA Pressure > 25 mm Hg | 29,638 (75.0) | 1,616 (76.4) | 1,445 (70.0) | N/A |
| PA Pressure ≤ 25 mm Hg | 9,895 (25.0) | 500 (23.6) | 622 (30.0) | N/A |
| **Mitral Regurgitation** | 26,412 (48.6) | 1,438 (49.6) | 1028 (42.6) | 76 (4.0) * |
| **Aortic Regurgitation** | 4,602 (8.5) | 232 (8.0) | 144 (6.0) | 29 (1.5) * |
| **Aortic Stenosis** | 3,668 (6.8) | 218 (7.5) | 155 (6.4) | 209 (11.0) * |
| **Difficult Study** | 7,566 (13.8) | 446 (15.3) | 179 (7.4) | N/A |

\*1916 studies in the SHC test cohort had information on Demographics and EF. 1897 studies in the SHC test cohort that had information on comorbidities.

### View Classifier Performance Across Two Institutions

On a test set of 2,462 TTEs (101,415 videos) from CSMC not seen during model training, the view classifier had an AUC of 1.000 (0.999-1.000) and at a threshold of 0.800, identified an A4C video with colour doppler across the tricuspid valve in 2,410 studies in the test set with a sensitivity of 0.979 (0.973-0.985) and a specificity of 1.000 (1.000 – 1.000). To evaluate generalization of the view classification model at a geographically distinct site, we evaluated its performance on 5,549 studies (278,377 videos) from SHC. In this external validation set, the view classifier picked up at least one A4C tricuspid Doppler video in 5,268 studies for a sensitivity of 0.949 (0.944 - 0.955) and a specificity of 1.000 (0.999 – 1.000)

### Tricuspid Regurgitation Severity Performance Across Two Institutions

The TR severity model showed strong performance in TR detection (**Figure 2**). In the temporally distinct CSMC test set, the model detected at least moderate TR with an AUC of 0.928 (0.913 - 0.943) and detected severe TR with AUC of 0.956 (0.940 - 0.969). Severe TR was ruled out with an NPV of 0.966 (0.955 - 0.977) and at least moderate TR was excluded with an NPV of 0.893 (0.871 - 0.914). Further information on TR model performance is presented in **Table 3**. Strong model performance was preserved across institutions. In the 2018 SHC cohort, the model identified severe TR with an AUC of 0.980 (0.966 – 0.989) and moderate/severe TR (defined as at least moderate TR) with an AUC of 0.951 (0.938 - 0.962). In this cohort, the model demonstrated an NPV of 0.987 (0.982 – 0.991) for severe TR and an NPV of 0.994 (0.990 – 0.997) for ≥ moderate TR.

**Figure 2:**
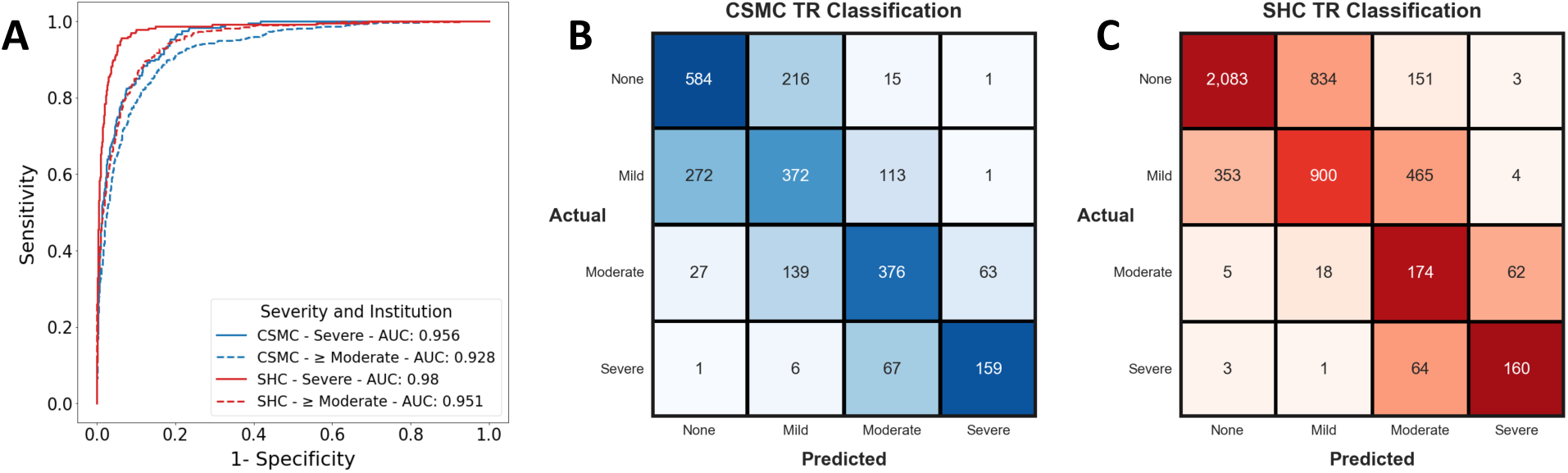
Model Performance Across Severity and Institution. - A. Receiver operating characteristic (ROC) curves for detection of Severe or ≥ Moderate TR at CSMC and Stanford. “> Moderate” included moderate, moderate to severe, and severe TR. 3B and 3C: TR Classification on test set videos from CSMC and Stanford, respectively. Confusion matrix colourmap values were scaled based on the proportion of actual disease cases in each class that were predicted in each possible disease category. This was done to allow for relative comparison of model performance across disease classes (None, Mild, Moderate, and Severe) given class imbalance. Statistics and confusion matrices are reported on a study level. When a study had multiple TR Doppler videos, the video with the max predicted TR severity was used. When multiple videos lead to the same maximum severity model prediction, the video with the highest prediction probability for that severity of TR was used.

**Table 3.** Model Performance.

| **<u>Site</u>** | **<u>Class</u>** | **<u>AUROC</u>** | **<u>PPV</u>** | **<u>NPV</u>** | **<u>Recall</u>** | **<u>F1-Score</u>** |
| --- | --- | --- | --- | --- | --- | --- |
| CSMC | ≥ Moderate TR | 0.928<br>(0.913 - 0.943) | 0.836<br>(0.799 - 0.871) | 0.893<br>(0.871 - 0.914) | 0.794<br>(0.753 - 0.831) | 0.814<br>(0.784 - 0.843) |
|  | Severe TR | 0.956<br>(0.940 - 0.969) | 0.710<br>(0.622 - 0.791) | 0.966<br>(0.955 - 0.977) | 0.682<br>(0.596 - 0.765) | 0.696<br>(0.622 - 0.759) |
| Stanford | ≥ Moderate TR | 0.951<br>(0.938 - 0.962) | 0.427<br>(0.383 - 0.466) | 0.994<br>(0.990 - 0.997) | 0.945<br>(0.914-0.971) | 0.586<br>(0.544 - 0.626) |
|  | Severe TR | 0.980<br>(0.966 - 0.989) | 0.699<br>(0.614 - 0.780) | 0.987<br>(0.982 - 0.991) | 0.702<br>(0.616 - 0.784) | 0.700<br>(0.630 - 0.763) |

### Subset Analysis

The TR severity model showed strong performance across test set subgroups **(Table 4)**. In studies with normal, mildly depressed, or moderately/severely depressed RV function, moderate/severe TR was detected with AUC of 0.923 (0.902-0.942), 0.904 (0.847 – 0.951), and 0.861 (0.848 – 0.952) respectively. Meanwhile severe TR was identified with AUC of 0.962 (0.940 – 0.979), 0.924 (0.872 – 0.966), and 0.882 (0.873 – 0.966). Performance was similar across different LVEF ranges. The model also performed similarly well in studies from patients with a history of pre-existing atrial fibrillation, right atrial dilation, and co-existing left sided valvular heart disease, with moderate/severe TR detection ranging from 0.886 (0.804 – 0.954) to 0.918 (0.848 – 0.973) and severe TR detection ranging from 0.917 (0.975 – 0.952) to 0.961 (0.908 – 0.997).

**Table 4.** TR Model Subset Analysis.

| Subset | Number of Studies | TR Severity Model Performance |
| --- | --- | --- |
| **RV Function** |  |  |
| Normal Function | 1845 | Moderate/Severe: 0.923 (0.902 - 0.942)<br>Severe: 0.962 (0.940 - 0.979) |
| Mildly Depressed | 285 | Moderate/Severe: 0.904 (0.847-0.951)<br>Severe: 0.924 (0.872 - 0.966) |
| Moderately/Severely Depressed | 266 | Moderate/Severe: 0.861 (0.848 - 0.952)<br>Severe: 0.882 (0.873 - 0.966) |
| **LV Systolic Function** |  |  |
| LVEF ≥ 50 | 1924 | Moderate/Severe: 0.930 (0.913 – 0.947)<br>Severe: 0.962 (0.944 – 0.977) |
| LVEF < 50 | 436 | Moderate/Severe: 0.897 (0.851 – 0.937)<br>Severe: 0.920 (0.876 – 0.957) |
| LVEF ≥ 35 | 2105 | Moderate/Severe: 0.932 (0.915 – 0.947)<br>Severe: 0.960 (0.943 – 0.974) |
| LVEF < 35 | 255 | Moderate/Severe: 0.860 (0.787 – 0.923)<br>Severe: 0.917 (0.857 – 0.964) |
| **Atrial fibrillation** | 499 | Moderate/Severe: 0.908 (0.869 - 0.941)<br>Severe: 0.917 (0.875 – 0.952) |
| **RA Dilation** | 505 | Moderate/Severe: 0.892 (0.847 – 0.931)<br>Severe: 0.920 (0.882 – 0.952) |
| **Pulmonary Artery Pressure** |  |  |
| PA Pressure > 25 mm Hg | 1,445 | Moderate/Severe: 0.892 (0.871 – 0.916)<br>Severe: 0.929 (0.905 – 0.950) |
| PA Pressure ≤ 25 mm Hg | 622 | Moderate/Severe: 0.920 (0.835 – 0.982)<br>Severe: 0.998 (0.990 - 1.000) |
| PA Pressure > 35 mm Hg | 822 | Moderate/Severe: 0.850 (0.806 – 0.890)<br>Severe: 0.881 (0.841 – 0.916) |
| PA Pressure ≤ 35 mm Hg | 1,245 | Moderate/Severe: 0.915 (0.880 – 0.945)<br>Severe: 0.990 (0.975 - 0.999) |
| **Mitral Regurgitation** | 357 | Moderate/Severe: 0.896 (0.869 - 0.922)<br>Severe: 0.945 (0.920 – 0.966) |
| **Aortic Stenosis** | 155 | Moderate/Severe: 0.918 (0.848 - 0.973)<br>Severe: 0.959 (0.901 - 0.996) |
| **Aortic Regurgitation** | 144 | Moderate/Severe: 0.886 (0.804 - 0.954)<br>Severe: 0.961 (0.908-0.997) |
| **Study Quality** | 253 | Moderate/Severe: 0.927 (0.826 - 0.989)<br>Severe: 0.982 (0.932 – 1.000) |

### MRI Comparison

Model predictions of TR severity were compared with assessments using cardiac magnetic resonance (CMR) imaging, another imaging modality capable of assessing valvular function. The MRI comparison cohort was composed of 748 studies from 572 patients. The cohort consisted of 468 studies (62.6%) with no TR, 243 studies (32.5%) with mild TR, 25 studies (3.3%) with moderate TR, and 12 studies (1.6%) with severe TR **(Supplemental Table 1).** Model predicted TR severity showed strong concordance with CMR assessment of TR severity for moderate/severe TR (AUC: 0.896 (0.822 – 0.948)) and severe TR (AUC: 0.949 (0.845 – 0.999)) (**Supplemental Table 2**). Similarly, cardiologist-determined TR severity from echo also agreed with cardiologist-determined severity using MRI for moderate/severe TR 0.820 (0.686 – 0.966) and severe TR 0.841 (0.480 – 0.997) (Supplemental Table 3). Meanwhile, the difference in AUC between AI model predictions and cardiologist-based echo prediction vs. MRI were not significantly different for at least moderate TR (DeLong test, p = 0.11) or severe TR (DeLong test, p = 0.08) (**Supplemental Table 4**). Full results are shown in **Supplemental Figure 1**.

### Model Explainability

The Integrated Gradients method was used to create saliency maps that identified regions of interest in each video contributing the most to detection of TR severity (**Figure 3**).^29^ Saliency maps for the TR severity model demonstrated that the clinically relevant imaging features of TR were important for model predictions, with activation signal localizing to pixels in the colour Doppler window and primarily highlighting the TR jet, indicating that the model used appropriate, physiologic features of TR to make predictions. **Supplementary Videos S1-S4** contain frame-by-frame visualizations of saliency maps.

**Figure 3:**
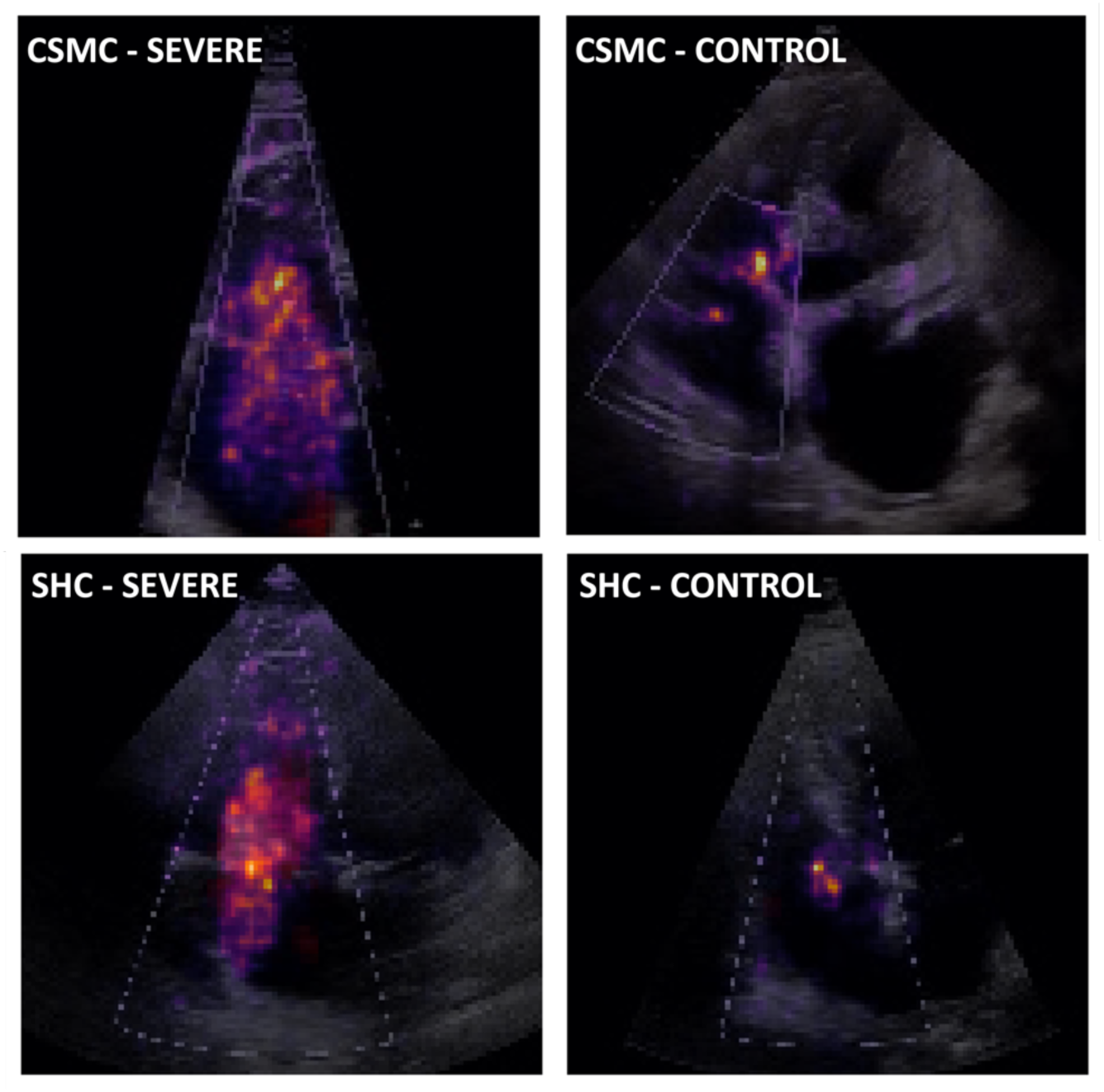
Saliency Map Visualization for TR classification models -. Videos with severe TR from CSMC (top left) and SHC (bottom left) and videos with no TR from CSMC (top right) and SHC (bottom right) are included. Saliency maps were computed using the Integrated Gradients method. The maximum value along the temporal axis for each pixel location was used to generate the final 2-dimensional heatmap. In the colourmap, pixels more salient to model predictions are brighter in colour and closer to yellow. Pixels darker in colour are less influential to the model’s final prediction. Severe TR videos were assessed by using the activation function for severe TR output neuron to generate a heatmap. When assessing controls (cases with no TR), heatmaps were generated by stacking heatmaps for severe and moderate TR output neurons and taking the maximum between the two at each pixel location.

## Discussion

The current work presents a comprehensive, automated pipeline capable of characterizing tricuspid regurgitation from echocardiogram studies. From full studies, the algorithm automatically selects A4C videos with colour Doppler across the tricuspid valve and assesses TR severity. In a temporally distinct test cohort, the pipeline demonstrated strong performance in severe and ≥ moderate TR screening, with AUC > 0.928 and NPV > 0.893. Moreover, this automated workflow robustly generalized to thousands of studies from a large, geographically distinct cohort. Given these characteristics, our results suggest a deep learning model could aid in the preliminary assessment of TR, facilitate review of institutional databases, or expand access for screening in low-resource settings.

In AI applied to echocardiography, the right heart is still under-represented. While prior work have shown AI model’s ability to characterize LVEF, LVH, and left sided valvular lesions, ^7,30,8^ there has been little work on TR. The present work applies a state-of-the-art video-based architecture for TR detection on a large training dataset to show such an approach produces a generalizable assessment of regurgitant severity. Despite relying on a single view, the performance of EchoNet-TR reliably matches expert cardiologists’ assessments across entire studies. TR assessment is a complex task, reliant multiple echocardiographic views, however the current pipeline proposed here performs well even when using the A4C view alone, suggesting a richness of ancillary information (right atrial enlargement, right ventricular systolic function, etc.) possibly related to TR severity. This potential use of this information mimics clinical decision making, where cardiologists often integrate conflicting metrics from different views and ancillary information to arrive at a final TR severity.

While the current work is promising, limitations should be considered. The regurgitant TR jet is 3-dimensional, and a combination of multiple echocardiographic views offers comprehensive visualization. Despite the strong performance, reliance on the A4C view alone could result in information loss that could still lead to incorrect predictions, especially if the TR is acute. Future work could focus on automatic quantification of valve leaflet thickness, orifice area, and other quantitative metrics of TR severity and risk stratification for tricuspid valve intervention. Similarly, with prior works using AI to guide novices to obtain videos of standard view echocardiogram views, the current algorithm could potentially be used with existing models to guide imaging acquisition to increase access to TR screening.^18,31^ Future work could focus on automatically quantifying parameters related to TR, high-throughput phenotyping aetiology based classification of TR, or precision medicine based stratification for intervention on the tricuspid valve.

In summary, we introduce a model for TR screening from single-view TTE videos. In doing so, we provide a workflow for isolating tricuspid valve colour Doppler videos and assessing TR severity with strong AUC and in two distinct test cohorts. Given its excellent performance and generalizability, the deep learning pipeline could aid point-of-care TR screening or enable retrospective institutional database review.

## Acknowledgements

At the time of part of this work, A.V. was a research fellow supported by the Sarnoff Cardiovascular Research Award.

## Funding

At the time of part of this work, A.V. was a research fellow supported by the Sarnoff Cardiovascular Research Award. D.O. reports research grants R00 HL157421 and R01HL173526 and support from AstraZeneca Alexion, as well as consulting from EchoIQ, Ultromics, Pfizer, and InVision.

## Disclosure of Interest

D.O. reports research grants R00 HL157421 and R01HL173526 and support from AstraZeneca Alexion, as well as consulting from EchoIQ, Ultromics, Pfizer, and InVision. A.C.K. reports support consulting fees from InVision.

## Data Availability

The dataset of videos and reports used to train and test EchoNet-TR is not publicly available due to its potentially identifiable nature.

## Code Statement

Our code and model weights are available at https://github.com/echonet/TR

## Notes

### Author Declarations

This study was approved by the Institutional Review Boards at Cedars-Sinai Medical Center and Stanford Healthcare. Informed consent was waived as the study involved secondary analysis of existing data without patient contact.

