## Supplemental Material for "Deep Learning Phenotyping of Tricuspid Regurgitation for Automated High Throughput Assessment of Transthoracic Echocardiography"

### Supplemental Figure 1

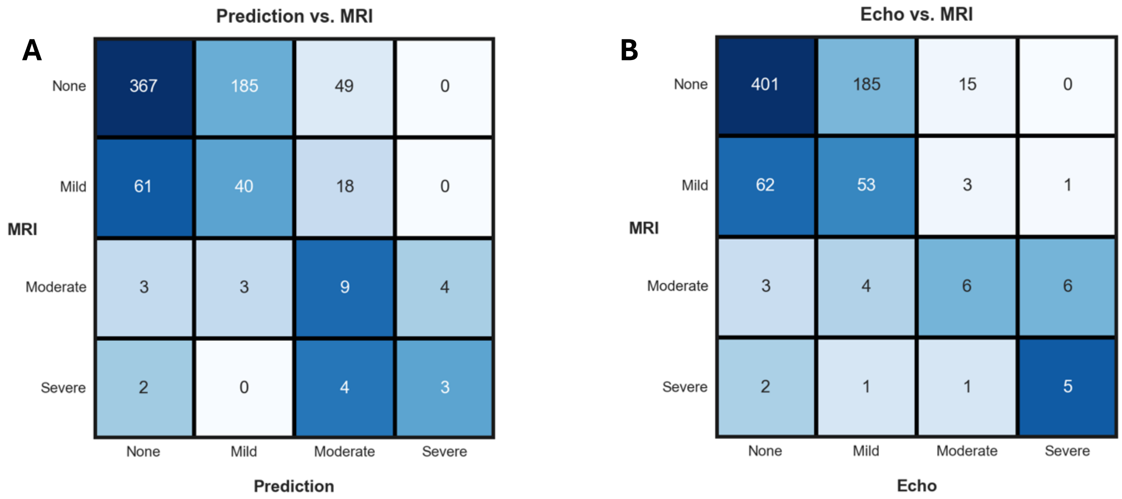


### MRI Cohort Characteristics

| | |
|---|---|
| **Number of Patients** | 572 |
| **Number of Studies** | 748 |
| **TR Severity** | |
| **None** | 468 (62.6) |
| **Mild** | 243 (32.5) |
| **Moderate** | 25 (3.3) |
| **Severe** | 12 (1.6) |

### Echocardiogram Severity vs. MRI Severity

| Severity | AUC |
|---|---|
| **Moderate/Severe** | 0.820 (0.686 – 0.966) |
| **Severe** | 0.841 (0.480 – 0.997) |

### Model Predicted Severity vs. MRI Severity

| Severity | AUC |
|---|---|
| **Moderate/Severe** | 0.896 (0.822 – 0.948) |
| **Severe** | 0.949 (0.845 – 0.999) |

### DeLong Test Results

| Severity | p-value |
|---|---|
| **Moderate/Severe** | 0.11 |
| **Severe** | 0.08 |
